# Hemoglobin-Albumin-Lymphocyte-Platelet (HALP) score as a predictive value of Incidental prostate cancer for patients going for Transurethral resection of the prostate (TURP): A Single Center Study

**DOI:** 10.1101/2024.01.23.24301670

**Authors:** Ahmed Bendari

## Abstract

**Aims:** Prostate cancer is a significant health concern worldwide, and early detection is crucial for effective treatment. This study aimed to investigate the role of the Hemoglobin-Albumin-Lymphocyte-Platelet (HALP) score in detecting prostate cancer in patients undergoing Transurethral Resection of the Prostate (TURP). Additionally, comprehensive analysis was performed to explore clinical parameters associated with incidentally diagnosed prostate cancer post-TURP.

**Methods:** A total of 131 patients with symptomatic bladder outlet obstruction who underwent TURP were included in the study. The patients were divided into two groups: those with benign prostatic hyperplasia (BPH) and those with invasive prostatic carcinoma. The IPC group consisted of patients with both low-grade and high-grade IPC determined by Gleason score. Demographic data, including age, race, medical history, body mass index, smoking and alcohol status, and family history of prostate cancer, were evaluated. Postoperative measurement of specimen weight and prostate-specific antigen (PSA) levels were also analyzed.

**Result:** Results revealed that approximately 50% of patients had BPH, while the remaining 50% had IPC. Patients with IPC, particularly high-grade IPC, had significantly higher PSA levels and lower resected specimen weight compared to those with BPH. The HALP score, which incorporates hemoglobin, albumin, lymphocyte, and platelet levels, showed promise as a discriminatory tool for distinguishing between BPH and IPC, as well as between high-grade IPC and BPH/low-grade IPC. Logistic regression analysis identified increased PSA levels, decreased HALP score, and smaller specimen weight as independent predictive factors for IPC after TURP. Notably, HALP score was the only significant independent predictive factor associated with high-grade IPC.

**Conclusion:** These findings contribute to the understanding of risk factors and diagnostic tools for incidentally detected prostate cancer in patients with bladder outlet obstruction undergoing TURP. The HALP score, along with PSA levels and specimen weight, can aid in the early detection and management of prostate cancer. Further research is warranted to validate these findings and explore the clinical utility of the HALP score in predicting prostate cancer outcomes.

## 1. Introduction

Prostate cancer is the second most frequent cancer diagnosis made in men and the fifth leading cause of death worldwide. Prostate cancer may be asymptomatic at an early stage and often has an indolent course.

Prostate cancer is graded based on Gleason score system which determines the aggressiveness of the prostate cancer. The score ranges from 2 to 10 and is calculated by adding the grades of the two largest areas of the cancer tissue. The lower the score, the more the cancer cells resemble healthy prostate cells. The higher the score, the more abnormal and likely to spread the cancer cells are A higher Gleason score is associated with a higher grade of cancer and likelihood of metastasis.

Inflammation and nutrition have been reported to be associated with cancer progression, drug response and cancer survival. Many inflammatory or nutritional prognostic indices, including the neutrophil-to-lymphocyte ratio (NLR), lymphocyte-to-monocyte ratio (LMR), prognostic nutrition index (PNI), and albumin-to-globulin ratio (AGR), have been developed to predict survival outcomes in solid tumors. The hemoglobin, albumin, lymphocyte, and platelet (HALP) score, a novel biomarker that was defined as hemoglobin × albumin × lymphocytes/platelets, was first introduced in 2015 by Chen *et al*. for predicting survival outcomes in patients with gastric cancer. A lower HALP score, which is formulated by hemoglobin × albumin × lymphocytes/platelets, is correlated with worse survival outcomes.

In our study we aim to evaluate the role of HALP score in detecting PC in patients undergoing TURP. Additionally, we also aim to make a broad comprehensive analysis of patients who are incidentally diagnosed of PC after TURP procedure, based on clinical parameters such as age, race, past medical history (diabetes, hypertension), body mass index, smoking and alcohol status and family history of prostate cancer, and postoperative measurement of specimen weight.

## 2. Methods

### 2.1. Study Design and Data Collection

This study retrospectively utilized our electronic medical record system (Cerner) at Lenox Hill Hospital. The study included all patients who underwent TURP between January 2019 and January 2023, meeting specific criteria such as having no significant medical conditions other than benign prostate hyperplasia and being incidentally diagnosed with prostate cancer after histologic examination. A total of 131 cases were included in the analysis. Patient data, including age, race, BMI, comorbidities, family history of prostate cancer, smoking and alcohol status, preoperative laboratory factors, and postoperative specimen weight, were collected from the hospital’s electronic medical record systems (Sunrise and Allscripts) and the pathology database system (Cerner).

### 2.2. Patient Characteristics

We searched our pathology system (Cerner) for keywords related to benign prostatic hyperplasia, transurethral resection of the prostate, and prostate chips. A total of 332 charts were extracted and manually checked for inclusion criteria, resulting in a final sample size of 131 cases. Exclusion criteria included prior history of prostate cancer or other malignancies, medical conditions impacting hemoglobin and platelet count, and conditions affecting albumin level, as well as cases with preoperative active infection or fever. Two pathologists have reviewed all cases, and PIN 4 and P63 IHC were performed on all equivocal cases.

### 2.3. Tumor grading and scoring and HALP score calculation

We categorized the cases into three groups: Benign prostatic hyperplasia, low grade prostate cancer and high-grade prostate cancer. Low grade group is defined as prostate cancer with Gleason score 6 (Gleason pattern = 3+3) and 7 (Gleason patterns = 3+4 or 4 + 3). And high-grade group is defined as prostate cancer with Gleason score 8 (Gleason patterns = 4+4 or 3+5 or 5+3), 9 (Gleason patterns = 5+4 or 4+5) and 10 (Gleason pattern = 5+5).

The HALP score is calculated as HALP Score = [hemoglobin (g/dL) × albumin (g/ dL) × absolute lymphocytes count (/uL)]/platelets (/uL)

### 2.4. Statistical Analysis

Data analysis was performed using Jjamovi (Version 2.3). Descriptive statistics such as median (interquartile range) and frequency and percentage were reported for continuous and categorical variables, respectively. Non-parametric tests, such as Kruskal-Wallis and chi-square tests, were used for analyzing continuous and categorical variables, respectively. Bivariate analysis was used to assess variable associations. A p-value of < 0.05 was considered statistically significant. Receiver operating characteristics (ROC) curves were used to determine sensitivity and specificity, and logistic regression models were used to calculate odds ratios and 95% confidence intervals for predictive factors. (*Reference: The Jamovi project (2022). Jamovi. (Version 2*.*3) [Computer Software]. Retrieved from* ***https://www.jamovi.org***.)

## 3. Results

In the current study, total participants were 131 patients with symptomatic bladder outlet obstruction who underwent TURP were included. The median (IQR) of age among participants was 71 (13) years.

BPH was detected in 66 patients (50.4%), while IPC was detected in the other 65 patients (49.6%). Fifty patients with IPC in TURP specimens classified as low-grade IPC according to Gleason score, and the other 15 patients classified as high-grade IPC. Demographic data of the patients were mentioned in (Table 1).

**Table 1.** Demographic data among studied patients (n=131)

| Variable | BPH<br>(n=66) | Low grade IPC<br>(n=50) | High grade IPC<br>(n=15) | P-value |
| --- | --- | --- | --- | --- |
| Age median (IQR) | 71 (11.5) | 70 (12.5) | 76 (12.5) | 0.2 |
| Age (N. %) |  |  |  |  |
| ≤71 years | 30 (45.5%) | 27 (54%) | 4 (26.7%) |  |
| >71 years | 36 (54.5%) | 23 (46%) | 11 (73.3%) | 0.2 |
| BMI median (IQR) | 27 (5.75) | 28 (7) | 27 (8.5) | 0.9 |
| BMI classification (N. %) |  |  |  |  |
| Underweight (BMI < 18.5) | 0 (0%) | 2 (40%) | 0 (0%) |  |
| Normal (18.5-24.9) | 17 (25.8%) | 12 (24%) | 3 (20%) |  |
| Overweight (25-29.9) | 29 (43.9%) | 19 (38%) | 6 (40%) | 0.7 |
| Obese (30 or higher) | 20 (30%) | 17 (34%) | 6 (40%) |  |
| Comorbidities |  |  |  |  |
| DM (N. %) | 48 (72.7%) | 11 (22.4%) | 2 (14.3%) | 0.6 |
| HTN (N. %) | 24 (36.4%) | 17 (34%) | 5 (33.3%) | 0.9 |
| History of smoking (N. %) | 32 (48.5%) | 28 (56%) | 10 (66.7%) | 0.4 |
| History of alcohol use (N. %) | 37 (56.1%) | 31 (62%) | 11 (73.3%) | 0.4 |
| Family history of prostate cancer | 8 (12.1%) | 11 (22%) | 3 (20%) | 0.4 |

\*Kruskal-Wallis test, Chi-Square test, Fisher-exact test

The IPC group had significantly higher PSA levels compared to the BPH group and among IPC group; patients with high grade IPC had higher PSA levels compared to low grade IPC patients (P=0.003), as most of patients with high grade IPC (46.7%) had PSA levels more than 10 ng/ml in comparison to (13.6% and 24%) in BPH and low-grade IPC patients respectively.

The weight of the resected specimens was significantly lower in high grade IPC patients in comparison to low grade IPC and BPH patients (P=0.03). A statistically significant difference was determined between low grade, high grade and BPH, with TUPR weight of 17 g taken as the cut-off point (P=0.04).

NLR, PLR and HALP scores were evaluated among the studied patients, there was statistically significant difference in NLR score (P=0.006) and highly statistically significant difference in HALP score (P<0.001) between the different studied groups (Table 2).

**Table 2.**
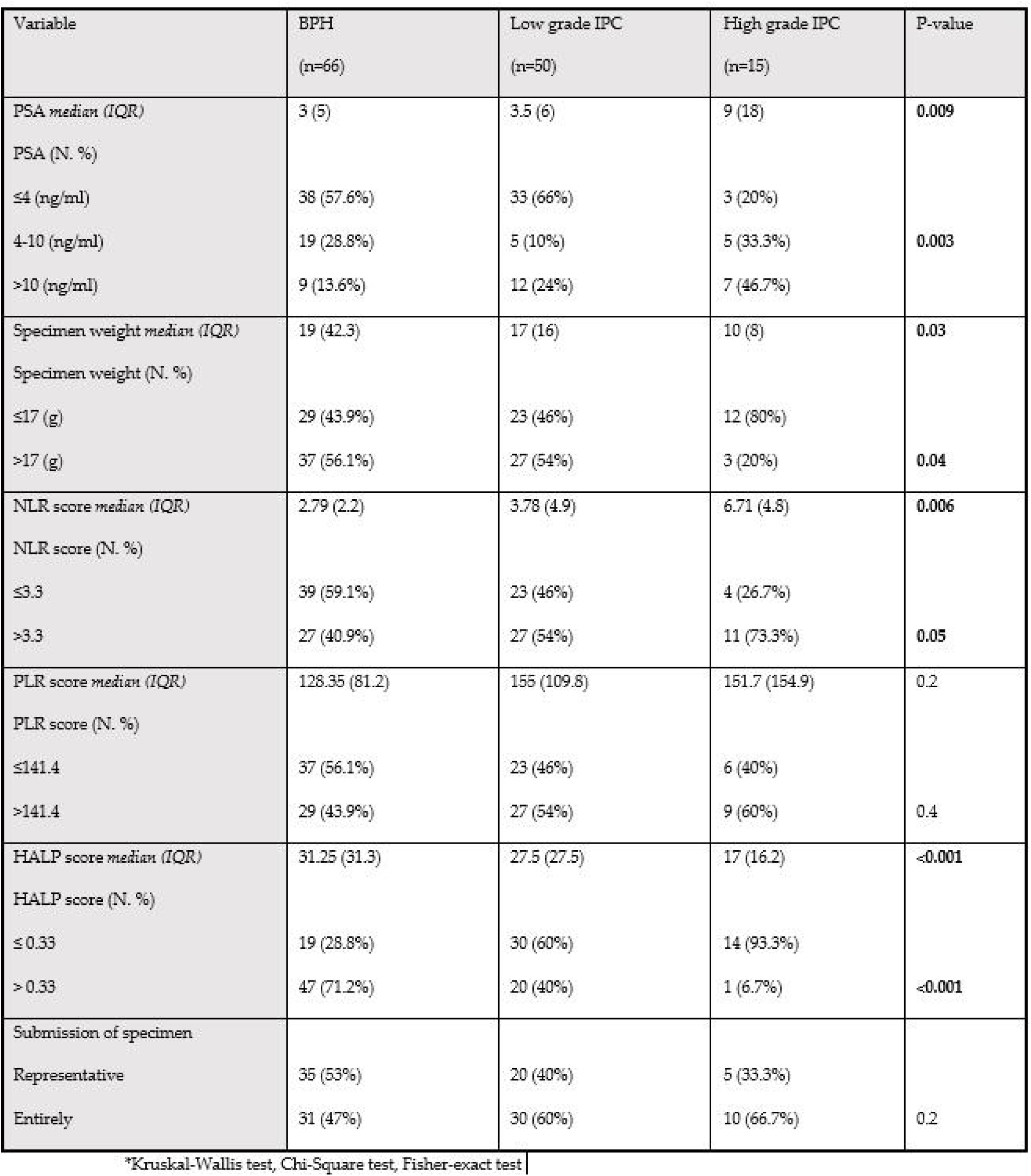
Characteristics among studied patients (n=131)

In ROC analysis, the AUC for discriminating patients with BPH from those with IPC according to HALP score was 0.738. According to the findings of the ROC analysis, the cut-off value for the HALP was set at 0.33. The sensitivity and specificity for the HALP score were 71.21% and 67.69% respectively. (Figure1)

**Figure 1.**
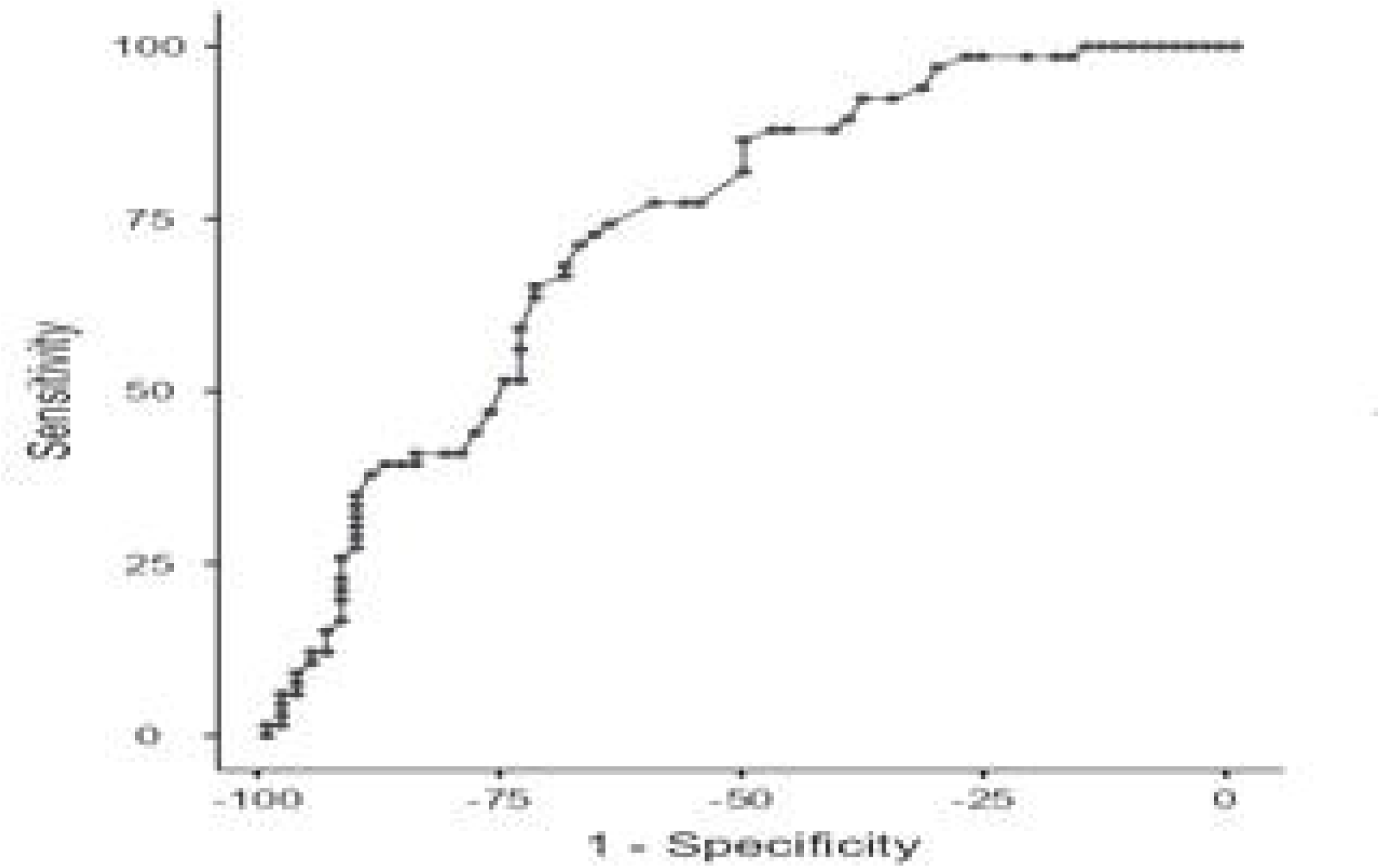
legend: Roc analysis to discriminate between BPH and IPC.

Another ROC analysis for discriminating patients with high grade IPC from BPH and low-grade IPC according to the HALP score was performed. The AUC was 0.8 with sensitivity 55.17% and specificity 93.33% at cut-off point 0.34, so HALP score could be considered as a good discriminator between patients with high grade IPC and those with BPH and low-grade IPC. (Figure2)

**Figure 2.**
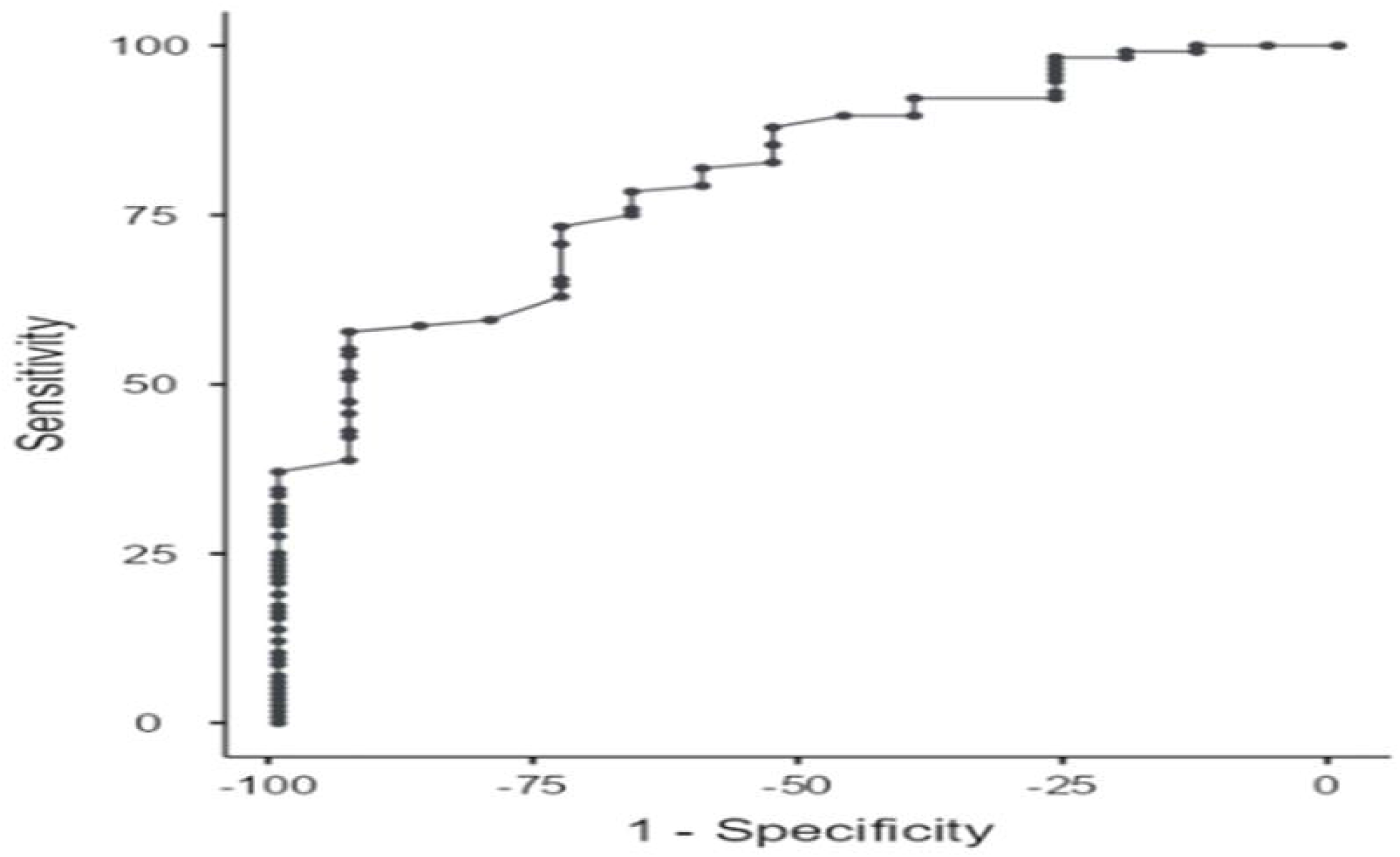
legend: ROC analysis of HALP score to discriminate high grade from low grade IPC and BPH.

To investigate the potential risk factors for IPC detection, we conducted logistic regression analyses using univariate and multivariate methods. In the univariate logistic regression, only PSA (odds ratio [OR], 0.03; 95% confidence interval [CI], 1.003–1.11; p=0.03), and HALP score (OR, 0.96; 95% CI, 0.94 – 0.98) were significantly associated with IPC after TURP, whereas no effect of other predictors was observed. Interestingly, the present study indicated that increased PSA (OR, 1.09; CI, 1.02–1.16) and decreased HALP score (OR, 1.01 with 95% CI, 0.002-0.13) were significant independent predictive factors for IPC after TURP. In addition, smaller specimen weight was associated with increased incidence of IPC (OR, 0.98 with 95 CI 0.97 – 0.99) after TURP. (Table 3)

**Table 3.** Logistic regression analysis for predictors of ICP.

| Variable | IPC |  |  |  |
| --- | --- | --- | --- | --- |
|  | Univariate |  | Multivariate |  |
|  | p-value | OR (95% CI) | p-value | OR (95% CI) |
| Age | 0.3 | 1.02 (0.98 – 1.06) | 0.2 | 1.03 (0.99 – 1.07) |
| PSA | 0.03 | 1.05 (1.00 – 1.11) | 0.02 | 1.09 (1.02 – 1.16) |
| NLR score | 0.45 | 1.01 (0.99 – 1.03) | - | - |
| PLR score | 0.16 | 1.00 (0.99 – 1.01) | - | - |
| HALP score | <0.001 | 0.96 (0.94 – 0.98) | <0.001 | 0.01 (0.002 – 0.13) |
| Specimen weight | 0.19 | 0.99 (0.98 – 1) | 0.007 | 0.98 (0.97 – 0.99) |

On conducting regression analysis for predictor factors of high-grade IPC, HALP score was the only significant independent predictive factor associated with high grade IPC (OR, 0.92 with 95% CI, 0.87 – 0.97). (Table 4)

**Table 4.** Logistic regression analysis for predictors of high-grade IPC.

| Variable | High grade IPC |  |  |  |
| --- | --- | --- | --- | --- |
|  | Univariate |  | Multivariate |  |
|  | p-value | OR (95% CI) | p-value | OR (95% CI) |
| Age | 0.07 | 1.06 (0.99 – 1.13) | 0.06 | 1.07 (0.99 – 1.15) |
| PSA | 0.08 | 1.01 (0.99 – 1.02) | 0.28 | 1.01 (0.99 – 1.02) |
| NLR score | 0.14 | 1.01 (0.99 – 1.03) | - | - |
| PLR score | 0.12 | 1.00 (0.99 – 1.02) | - | - |
| HALP score | <0.001 | 0.92 (0.87 – 0.96) | 0.004 | 0.92 (0.87 – 0.97) |
| Specimen weight | 0.053 | 0.95 (0.89 – 1.001) | 0.057 | 0.94 (0.88 – 1.00) |

## 4. Discussion

The global incidence of prostate cancer is on the rise. Current screening methods for prostate cancer involve a combination of digital rectal examination, elevated prostate-specific antigen (PSA) levels, and magnetic resonance imaging of the prostate. ^1^

However, these methods often yield false positive results in various benign conditions such as prostatitis and benign prostatic hyperplasia (BPH). Consequently, patients may undergo unnecessary biopsies or transurethral resection of the prostate (TURP), leading to unwarranted medical procedures, increased anxiety, and a heightened strain on healthcare systems.

Among these diagnostic methods, PSA testing garnered approval from the United States Food and Drug Administration (USFDA) as a screening tool for prostate cancer in 1994. Presently, a PSA cutoff level of 4 ng/mL is conventionally employed for screening purposes. The incorporation of PSA screening has notably contributed to a substantial escalation in the detection of prostate cancer.^2^ However, this advancement has concurrently resulted in an increased incidence of false-positive biopsies. This is attributed to the lack of specificity of PSA for prostate cancer, as elevated levels may be observed in benign conditions such as prostatitis and BPH. Furthermore, the limitations of PSA screening are evident in instances where PSA levels fall below the designated cutoff, resulting in missed detections in numerous cases of prostate cancer. Consequently, the exclusive reliance on PSA as a singular marker in screening practices has engendered a mixed perspective, owing to its susceptibility to false positives and the potential for false negatives.

In our investigation, we sought to assess the utility of the HALP score, which comprises hemoglobin, albumin, lymphocytes, and platelets, as a predictive tool for the occurrence of prostate cancer in patients undergoing TURP. Hemoglobin and albumin levels reflect nutritional status, while lymphocytes and platelets provide insights into the body’s immune status, all of which have known implications in the initiation and progression of cancer. The HALP score has primarily been employed to predict the prognosis of various malignancies, including gastric carcinomas, esophageal squamous cell carcinoma, colorectal cancer, renal cell carcinoma, bladder cancer, lung cancer, and pancreatic cancer.^3-8^ Furthermore, it has been explored as a prognostic indicator for metastatic prostate cancer post-radical prostatectomy, demonstrating its independence as a prognostic factor. ^9^Additionally, we computed two additional inflammatory markers, namely the neutrophil-to-lymphocyte ratio (NLR) and platelet-to-lymphocyte ratio (PLR) scores, recognized for their impact on overall survival outcomes in individuals with prostate cancer. Elevated NLR values have consistently been correlated with an unfavorable prognosis.^10-12^Analogously, heightened PLR values have been linked to diminished overall survival rates. ^13,14^

In a precedent investigation addressing the assessment of preoperative HALP scores among individuals diagnosed with oligometastatic prostate cancer subsequent to cytoreductive radical prostatectomy, a noteworthy correlation was established.^9^ Specifically, a diminished HALP score was significantly associated with a simultaneous reduction in progression-free. The utilization of HALP score threshold set at 32.4 was integral to this analysis. Contrastingly, an alternative study focused on the application of HALP scores in the diagnostic context of prostate cancer failed to elucidate significant distinctions in HALP scores between cases of BPH and prostate cancer.^15^ This particular investigation revealed higher HALP scores within the prostate cancer group compared to the BPH group, with median HALP scores of 51.2 and 49.43, respectively. These findings underscore the nuanced role of HALP scores and the need for context-specific considerations in different clinical scenarios.

Our investigation revealed that the HALP score is an independent marker for the detection of prostate cancer. Patients with prostate cancer exhibited lower HALP scores in comparison to those with BPH. Notably, at a HALP score of ≤ 0.33, the sensitivity and specificity for detecting prostate cancer were 71.2% and 67.69%, respectively. Additionally, our study indicated that the HALP score could effectively differentiate high-grade prostate cancer from low-grade prostate cancer, achieving a specificity of 93.33%.

We acknowledge that there were some limitations in the present study. The retrospective design from single center, and the small number of patients are the main limitations.

## 5. Conclusion

This study found that among patients with symptomatic bladder outlet obstruction who underwent TURP, approximately half had BPH while the other half had IPC. Patients with IPC, particularly those with high-grade IPC, had higher levels of PSA and lower resected specimen weight compared to patients with BPH. The HALP score showed promise as a discriminatory tool for distinguishing between BPH and IPC, as well as between high-grade IPC and BPH/low-grade IPC. Logistic regression analysis revealed that increased PSA levels, decreased HALP score, and smaller specimen weight were independent predictive factors for IPC after TURP. HALP score was the only significant independent predictive factor associated with high-grade IPC. These findings contribute to our understanding of risk factors and diagnostic tools for IPC in patients with bladder outlet obstruction.

## Data Availability

All data and materials are available upon request from the corresponding author

## Institutional Review Board Statement

I confirm that all relevant ethical guidelines have been followed, and any necessary IRB (Institutional Review Board) and/or ethics committee approvals have been obtained (the Feinstein Institutes for Medical Research/Northwell Health system located in New York City, USA). Ethical approval was waived; IRB approval number: 23-0717.

## Informed Consent Statement

Not applicable.

## Data Availability Statement

All data and materials are available upon request from the corresponding author.

